# Pilot study to evaluate hypercoagulation and inflammation using rotational thromboelastometry and calprotectin in COVID-19 patients

**DOI:** 10.1101/2022.05.27.22275701

**Authors:** S.N. Stanford, A. Roy, C. Rea, Ben Harris, A. Ashton, S. Mangles, T. Everington, R. Taher, D. Burns, E. Arbuthnot, T. Cecil

## Abstract

**Introduction:** Abnormal coagulation and inflammation are hallmarks of SARs-COV-19. Stratifying affected patients on admission to hospital may help identify those who are risk of developing severe disease early on. ROTEM is a point of care test that can be used to measure abnormal coagulation and calprotectin is a measure of inflammation.

**Aim:** Assess if ROTEM can measure hypercoagulability on admission and identify those who will develop severe disease early on. Assess if calprotectin can measure inflammation and if there is a correlation with ROTEM and calprotectin.

**Methods:** COVID-19 patients were recruited on admission and ROTEM testing was undertaken daily for a period of 7 days. Additionally inflammatory marker calprotectin was also tested.

**Results:** 33 patients were recruited to the study out of which 13 were admitted to ITU and 20 were treated on the ward. ROTEM detected a hypercoagulable state on admission but did not stratify between those admitted to a ward or escalated to ITU. Calprotectin levels were raised but there was no statistical difference (p=0.73) between groups. Significant correlations were observed between FIBA5 (p<0.00), FIBCFT (p<0.00), FIBMCF (p<0.00) and INMCF (p<0.00) and calprotectin.

**Conclusion:** COVID19 patients were hypercoagulable in admission. The correlations between ROTEM and calprotectin underline the interactions between inflammation and coagulation.

## Introduction

Patients infected with the coronavirus SARS-CoV-2 leading to the coronavirus disease 2019 (COVID-19) become symptomatic after an average incubation of 5.2 days (1). Severely affected patients develop shortness of breath at a median of 8 days from illness onset, with acute respiratory distress syndrome, pneumonia developing at day 9 and admission to Intensive Therapy Unit (ITU) at day 10.5 (2).

Biologically, severe COVID-19 is characterised by the production of proinflammatory cytokines (3). There is extensive cross talk between inflammation and coagulation systems in response to invasion by pathogens (4-6). Reflecting this, several studies have reported abnormalities in laboratory markers of coagulation and fibrinolysis in COVID-19 patients (7-11). Retrospective data indicates that there is significantly more derangement in coagulation parameters (namely prothrombin time (PT) and D-Dimer) at the point of admission in patients who don’t survive, versus those who do (12). These changes in individual coagulation parameters point to a disruption in haemostasis, but do not provide any guidance as to the biological effect of the changes. Traditional coagulation tests provide a snapshot of a particular aspect of coagulation in cell depleted plasma, but do not provide an assessment of overall haemostasis. Rotational thromboelastometry (ROTEM^®^) provides a composite assessment of the dynamic process of clot initiation, thrombin generation and whole blood clot formation which is arguably more representative of physiological processes (13).

A number of studies have used ROTEM^®^ to assess haemostatic status and demonstrate a prothrombotic phenotype in patients admitted to ITU (14-16). Furthermore, studies have also shown that ROTEM^®^ can be potentially used as a predictor for thrombosis and disease severity on admission (17-20).

In this study we investigated if COVID-19 patients demonstrated a prothrombotic phenotype as measured by ROTEM *Sigma* and if it could be used as a predictor of disease severity on admission to hospital. Correlations between ROTEM and inflammatory marker calprotectin were also undertaken to understand its role in contributing to hypercoagulation in COVID-19 patients.

## Materials and Methods

in line with the Declaration of Helsinki, ethical approval was obtained from the Cornwall and Plymouth Research Ethics Committee. IRAS No: 284755. As part of the ethical approval where patients had severe disease and were not able to consent, assent was obtained from an independent medical practitioner who was not involved in the direct care of the patient. Patients who were admitted to hospital with a suspected/known diagnosis of COVID-19 and over the age of 18 were included in the study. Patients who had a negative COVID-19 test after recruitment were subsequently excluded from analysis. Any patients with a previous history of coagulation disorders including venous thromboembolisn within 6 months and those on anticoagulant therapy excluding low molecular weight heparin thromboprophylaxis were excluded from the study. Admitted patients either went to a normal ward or to ITU depending on clinical severity. Patients with mild to moderate respiratory failure were administered oxygen via nasal cannula and venturi mask upto a maximum of 15 L/minute, if this was insufficient continuous positive airway pressure (CPAP) was administered. Those with severe respiratory failure were treated in ITU with invasive ventilation support. On admission patients were treated with a single prophylactic dose of enoxaparin in ED and then transferred to either a ward or ITU. Anticoagulation treatment was given twice daily (6am/6pm) and categorised based on weight. Enoxaparin 40mg if <100kg, 60mg if 100-150Kg and 80mg if >150kg. All patients were also given a stat dose of vitamin K IV with the first dose of enoxaparin. This was given as testing at the start of the pandemic had shown several patients had low protein C or S on admission.

Bloods were taken on admission and everyday for the first 7 days of hospital admission. The follow-up bloods were taken approximately 3 hours after anticoagulant treatment. Rotational thromboelastometry was tested within 4 hours of taking the blood and the plasma aliquots were frozen for batch testing of calprotectin.

### Rotational thromboelastometry

ROTEM *Sigma* (Werfen UK) is a viscoelastic point of care test with a fully automated system containing a sample handler and cartridge that uses whole blood to measure coagulation (21). All tests were analysed for 60 minutes and within 4 hours of sample collection. Reference ranges were obtained from healthy volunteer data (n=26) previously collected.

Data from three ROTEM channels are presented, FIBTEM, INTEM and EXTEM. FIBTEM uses cytochalasin D to inhibit platelet activity and provide a clot tracing that reflects the presence of fibrinogen. It provides information on fibrin formation and polymerisation without platelet contribution (22). In EXTEM tissue factor is used to initiate the extrinsic clotting cascade. The INTEM test uses ellagic acid to initiate clotting via the intrinsic pathway. The EXTEM and INTEM tests provide information on the extrinsic and intrinsic pathways respectively. The viscoelastic characteristics of the clot are measured from clot formation to lysis and include clotting time (CT), clot formation time (CFT), maximum clot firmess (MCF), A5-A30 (measure of clot firmess from 5-30 minutes) and maximum lysis (ML) which is the degree of fibrinolysis relative to MCF achieved during measurement and is reported as percentage of clot firmness lost (23). A shortened CFT with an increased MCF is indicative of a hypercoagulable state.

### Calprotectin

Circulating calprotectin levels were measured in plasma using a chemiluminescent immunoassay (QUANTA Flash, Werfen, UK) according to manufacturer’s instructions. The chemiluminescent immunoassay utilises a predefined lot specific master curve that is stored in the reagent cartridge barcode.

### Statistics

Demographics of the patient population are presented as frequencies and percentages. All continuous variables are presented with medians and interquartile ranges (IQR). Mann-Whitney U test was used to test whether there were significant differences between patients admitted to ITU vs ward for ROTEM and calprotectin. We included individuals in the ITU cohort if they were admitted to the high dependency unit (HDU) or intensive treatment unit (ITU) at any point during admission (n=13). The ward cohort represents the remaining individuals in the study (n=20). Spearman’s rank correlation coefficients were calculated to assess associations between variables. A Bonferroni correction was used to evaluate significance with both tests. A p-value below 0.05 at a 95% confidence interval was considered significant. Analysis was performed in Anaconda 3 with Python 3.8.8.

## Results

Blood was obtained from 47 patients admitted to Hampshire Hospitals from October 2020-April 2021. 14 patients were excluded as they tested negative for COVID-19, 33 who tested positive were included in the study. Rotational thromboelastometry demonstrated a hypercoagulable state compared to healthy controls, demographic data are presented in table 1. On admission for FIBTEM CT (p<0.001) and MCF (p<0.001), EXTEM CT, CFT, MCF and ML (p<0.002) and INTEM CT (p<0.028), MCF (p<0.004) and ML (p<0.001).

**Table 1:** Demography of patients admitted to hospital with COVID-19.

|  |  |  |
| --- | --- | --- |
| **Gender** | Female | 14 (42.4%) |
|  | Male | 19 (57.8%) |
| **Age** | (Median (IQR)) | 52 years (41.0 - 62.5) |
| **Smoking Status** | Yes | 2 (6%) |
|  | No | 24(72%) |
|  | Vape | 2 (6%) |
|  | Ex-smoker | 5 (21.21%) |
| **Comorbidities** | Diabetes | 9 (27.3%) |
|  | Hypertension | 8 (24.2%) |
|  | Cardiovascular | 6 (18.2%) |
|  | Cerebrovascular | 1 (3%) |
|  | Asthma/COPD | 6 (18.2%) |
|  | Malignancy | 1 (3%) |
|  | Other diseases | 15 (45.5%) |
|  | Nil | 6 (18.2%) |
| **COVID-19 diagnosed** | On admission | 25 (75.8%) |
|  | Later in ward | 8 (24.2%) |
| **Admission Unit** | Ward | 20 (60.6%) |
|  | ITU | 13 (39.4%) |
| **Length of Stay** | (Median (IQR)) | 8 days (5 - 16) |
| **Outcome / Event** | VTE | 1 (3.0%) |
|  | Discharge home | 29 (88.0%) |
|  | Deceased | 4 (12.0%) |

### ITU vs Ward

Comparison was made between ITU and wards patients, median values on admission were compared between the two groups. No Significant differences were seen between those admitted to ITU versus those admitted to the ward for any of the ROTEM parameters, table 2. Similarly, no significant differences were observed in calprotectin between the two groups. Longitudinal data for all ROTEM tests at the various time points for both ITU and ward cohorts are presented in Appendix 1.

**Table 2:** ROTEM parameters between those admitted to ward and ITU. The median values on admission are presented in the table.

| **ROTEM parameters** | **Ward (n=20)**<br>**Median (IQR)** | **ITU (n=13)**<br>**Median (IQR)** | **p value** |
| --- | --- | --- | --- |
| FIBCT (s) | 70 (65-77) | 67 (66-74) | 0.34 |
| FIBMCF (mm) | 28 (25-33) | 26 (24-30) | 0.15 |
| EXCT (s) | 70 (60-73) | 67 (59-71) | 0.31 |
| EXCFT (s) | 46 (43-57) | 57 (54-61) | 0.97 |
| EXMCF (mm) | 71 (68-74) | 72 (69-73) | 0.62 |
| EXML (%) | 5 (3-10) | 4 (2-6) | 0.06 |
| INCT (s) | 184 (162-197) | 185 (176-194) | 0.50 |
| INCFT (s) | 55 (49-71) | 58 (55-70) | 0.83 |
| INMCF (mm) | 68 (66-70) | 67 (66-70) | 0.51 |
| INML (%) | 5 (2-8) | 4 (3-6) | 0.23 |

Although not statistically significant (p=0.73) the calprotectin values were higher in the ITU patients; [median: 4.17; IQR:2.17-8.52] compared to those on the ward; [median:2.08; IQR: 1.11-3.53] the trends are in Figure 1. Averages for each time point are presented. Significant correlations were observed between FIBTEM, EXTEM and INTEM parameters and calprotectin, Table 3.

**Figure 1:**
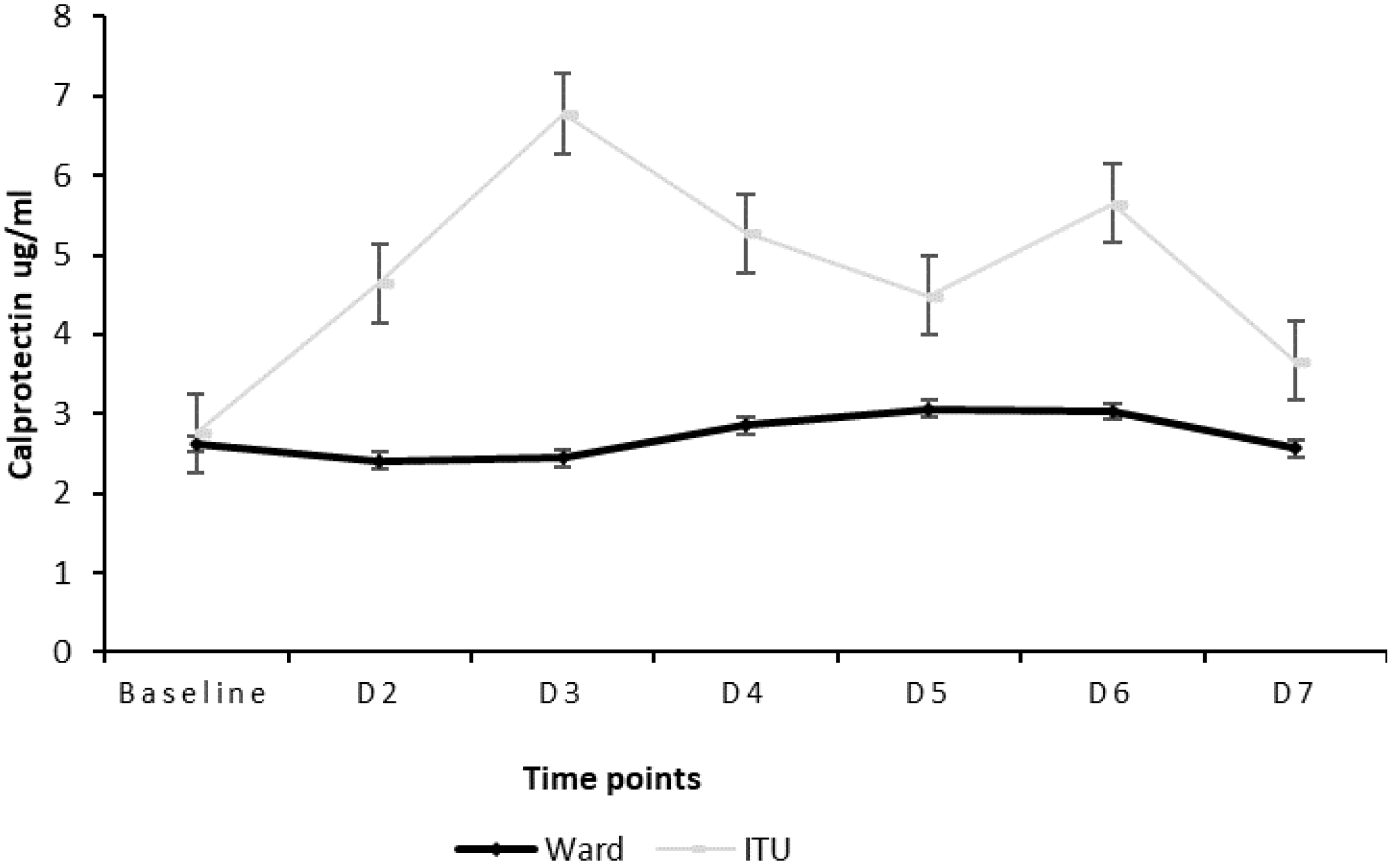
Calprotectin trend between ITU patients and those on the ward for admission through to week 2. Normal range for calprotectin is <1.99μg/ml.

**Table 3:** Spearman rank correlations between ROTEM and coagulation and calprotectin.

| ROTEM parameters | Calprotectin<br>r (p value) |
| --- | --- |
| FIBA5 | 0.62 (0.00)*** |
| FIBCFT | -0.57 (0.00)*** |
| FIBMCF | 0.64 (0.00)*** |
| EXA5 | 0.43 (0.01)** |
| EXMCF | 0.46 (0.01)** |
| INCT | 0.07 (0.68) |
| INA5 | 0.41 (0.02)* |
| INCFT | -0.41 (0.01)** |
| INMCF | 0.57 (0.00)*** |
| INLI30 | 0.1 (0.56) |

\* p-value less than 0.05
\*\* p-value less than 0.01
\*\*\* p-value less than 0.001 (significant after Bonferroni correction)

## Discussion

This pilot study confirmed hypercoagulability in COVID-19 patients on admission based on ROTEM parameters. However, ROTEM was not able to stratify on admission those who were escalated to ITU and those who were not.

This is in contrast with other studies that have also assessed if ROTEM can be used as a predictor of disease severity on admission. Almskog et al undertook ROTEM testing in COVID-19 patients who were admitted to regular wards and specialised ventilation support. All hospitalised patients with COVID-19 patients showed elevated values of EXMCF and FIBMCF on admission to hospital and this hypercoagulable pattern was more prominent in the severely ill patients compared to those on regular wards (17). Similarly, Boscolo et al investigated the difference in MCF values between those patients admitted to internal medicine wards and ICU. They found that ICU patients had a significantly higher FIBMCF compared to those admitted to the internal medicine wards (19). Lastly, ROTEM parameters were also measured in COVID-19 patients with varying severities of pneumonia; a hypercoagulable ROTEM pattern due to shortened EXCT, higher than normal EXMCF and FIBMCF and shortened EXCFT, high clot strength and hypofibrinolysis in advanced disease and patients with high levels of IL-6 were observed (20). It is likely that we did not see a difference between the two groups because of the small numbers of patients in our ITU cohort.

Calprotectin levels have been shown to be elevated in COVID-19 patients (24-26). In our study we found that calprotectin levels were raised overall, and patients admitted to ITU although not statistically significant had higher calprotectin levels compared to those admitted to a ward. Various studies have reported that raised calprotectin levels in COVID-19 patients. A marked difference in the serum calprotectin levels between COVID-19 survivors and non-survivors (24). Another study demonstrated that reduced frequency of non-classical monocytes along with raised serum calprotectin levels has the potential to identify patients who will develop severe COVID-19 (27). A review of calprotectin in COVID-19 provided evidence suggesting that calprotectin could be useful in assessing disease severity (28). Similar to our ROTEM results, it is likely we did not see a statistical difference between the two groups because of the small numbers in the ITU cohort.

We are the first to report significant correlations between FIBA5, FIBCFT and FIBMCF and calprotectin. It is known that fibrinogen is an acute phase reactant which is increased during an inflammatory response (29). In COVID-19 patients increased fibrinogen levels have been associated with excessive inflammation (30). We suggest that the significant correlation we have observed between fibrinogen and calprotectin is because of fibrinogen’s role in inflammation. Calprotectin also correlated with INMCF which measures the intrinsic pathway of coagulation. FVIII and VWF which are part of the intrinsic pathway of coagulation are also linked to inflammation (31-33) and are known to be raised in COVID-19 patients (33-37). Similar to fibrinogen, the link that FVIII and vWF have to inflammation could be the reason for the significant correlation calprotectin and INMCF. These interesting findings further highlights the role of inflammation in COVID-19 and its impact on coagulation which is measurable using ROTEM and should be investigated further. In addition to undertaking ROTEM testing in COVID-19 patients, consideration should be given to measuring calprotectin levels. ROTEM is a point of care test with quick turnaround times that gives an overview of coagulation. Calprotectin has the potential to be used as a marker of inflammation in these patients.

In summary, ROTEM detected a hypercoagulable state in COVID-19 patients but could not stratify severity of disease on admission in this pilot study. Significant correlations were observed between ROTEM and calprotectin demonstrating the synergy that exists between inflammation and coagulation.

### Limitations

No definite conclusions can be drawn from this pilot study because of the small sample size. The study consisted mainly of Caucasian patients and as coagulation status and predisposition to the development of coagulopathies varies between race and ethnicity (38) this limits the generalisability of our study.

### Addendum

Dr S. Stanford designed the study, collected the data and wrote the paper. Dr D. Burns, Ms R. Taher and Dr S. Stanford undertook statistical analysis. Ms E. Arbuthnot co-ordinated the study. All other authors reviewed and provided expert comments on the paper.

## Data Availability

ata cannot be shared publicly because of patient confidentiality. Data are available from the Hampshire Hospitals Institutional Data Access / Ethics Committee (contact via Sophia Stanford) for researchers who meet the criteria for access to confidential data.

## Acknowledgements

We would like to thank all our patients for participating in the study. The Peritoneal Malignancy research team-Natasha Beacher, Nicola Preston and Pennie Porter for all their hard work in recruiting the patients and undertaking ROTEM testing. Myk Saas for undertaking calprotectin testing and Werfen UK for providing the kits. This study was funded by the Peritoneal Malignancy Institute, kits for calprotectin testing were provided by Werfen, UK and kits.

## Appendix 1

Comparison of ITU vs ward for the various tests over time.

**Figure 1:**
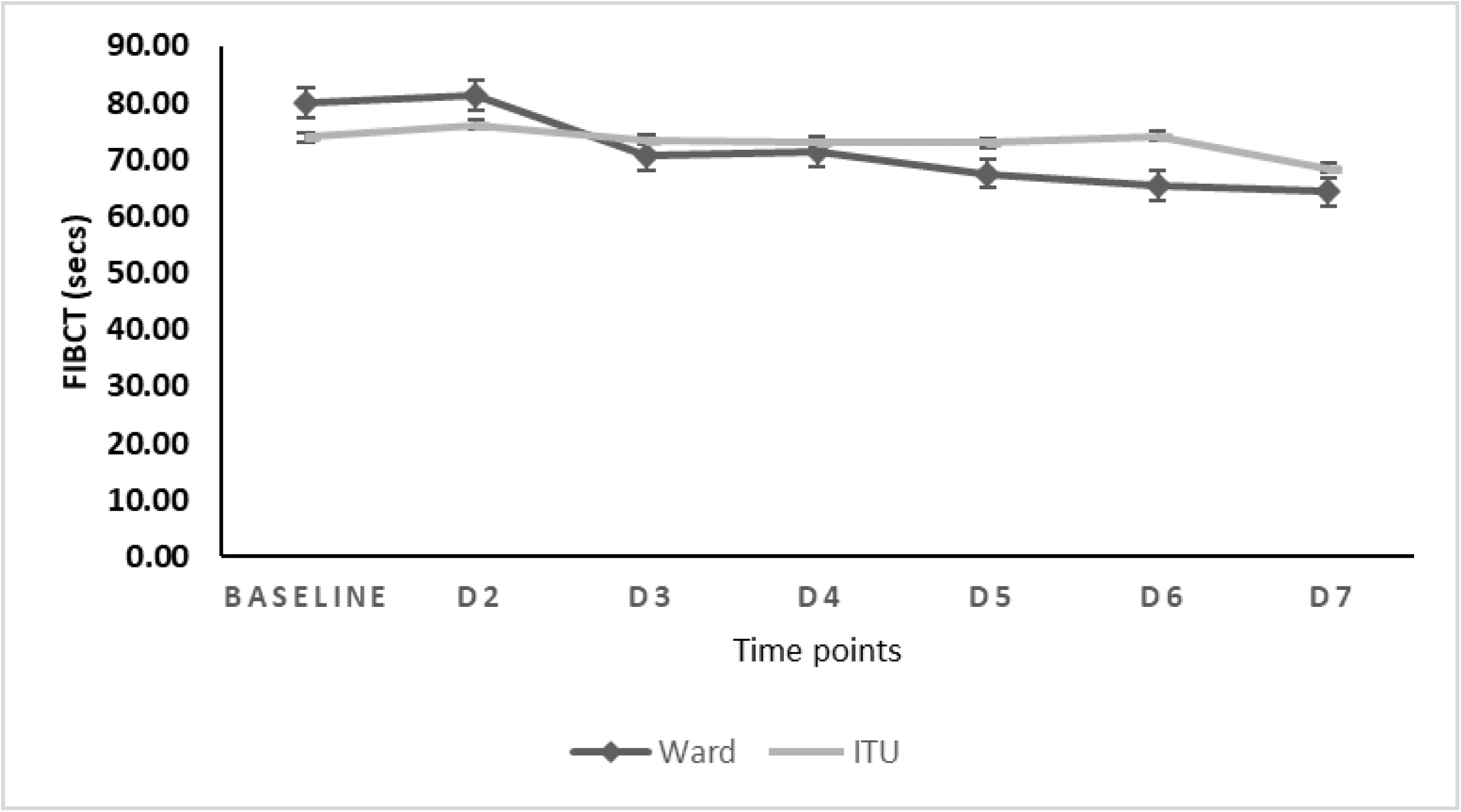
Values of FIBCT for COVID-19 patients over the first 7 days of admission to hospital. Comparison of FIBCT values between ITU and ward admissions.

**Figure 2:**
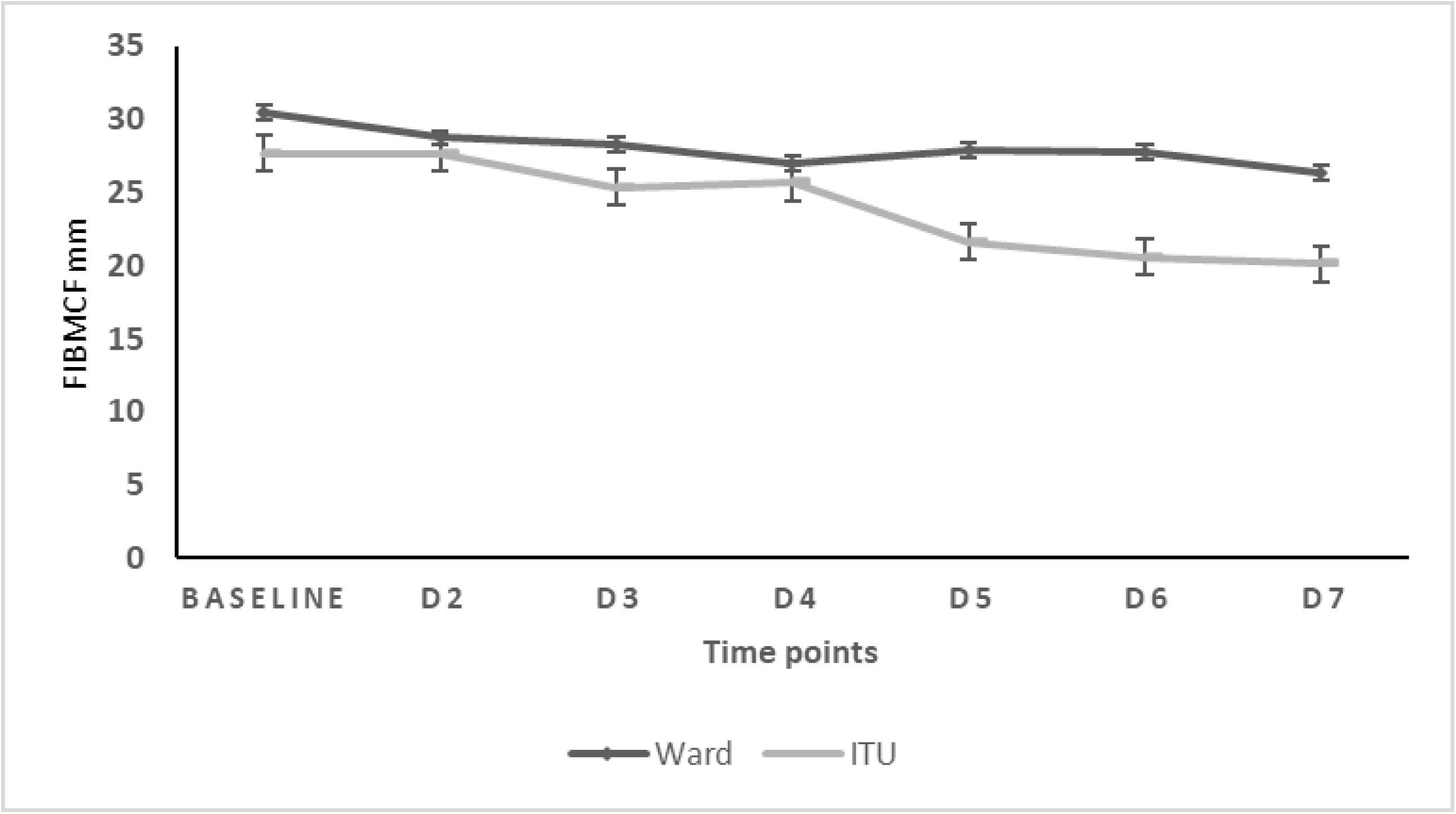
Values of FIBMCF for COVID-19 patients over the first 7 days of admission to hospital. Comparison of FIBMCF values between ITU and ward admissions.

**Figure 3:**
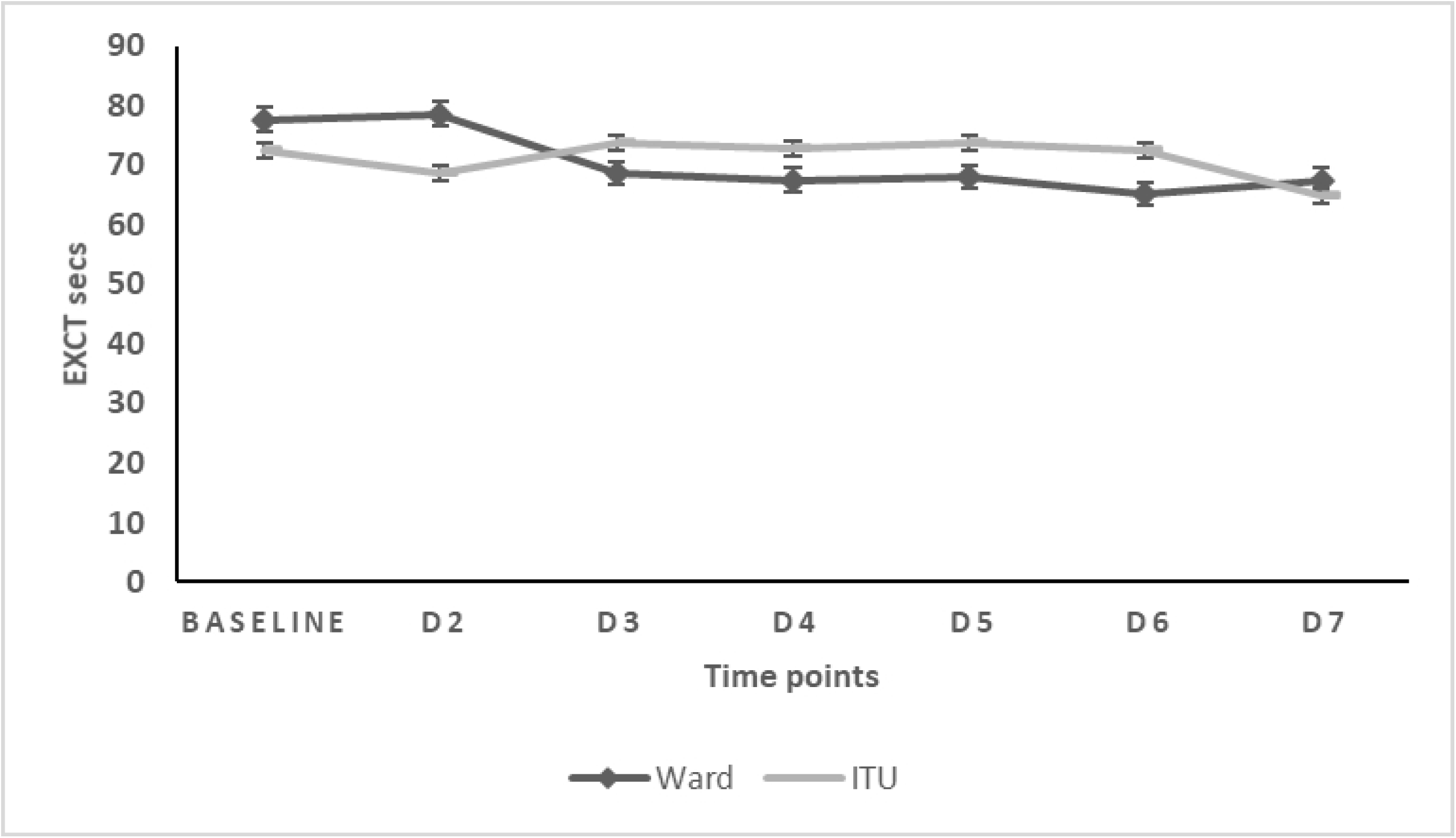
Values of EXCT for COVID-19 patients over the first 7 days of hospital admission. Comparison of EXCT values between ITU and ward admissions.

**Figure 4:**
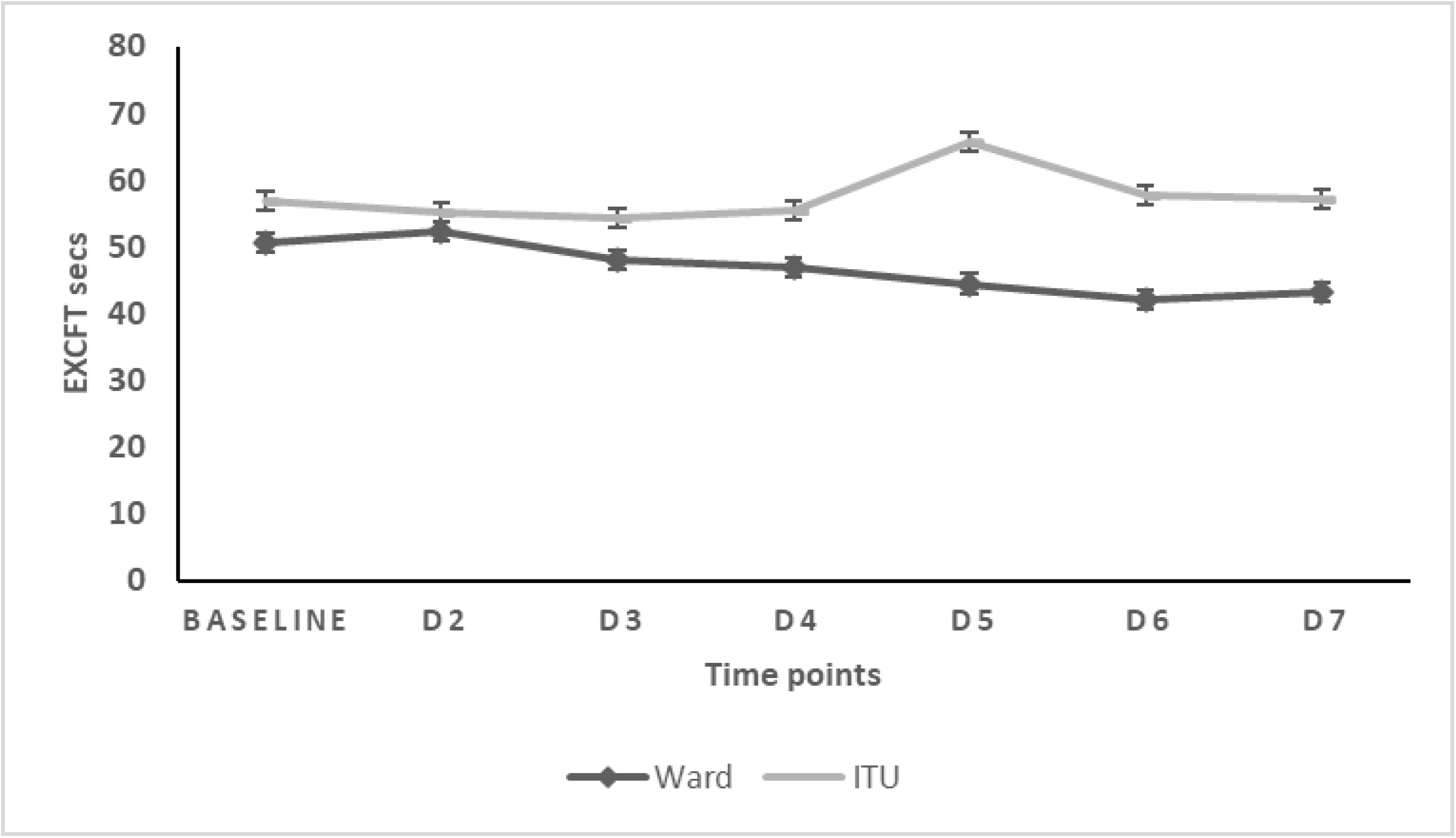
Values of EXCFT for COVID-19 patients over the first 7 days of hospital admission. Comparison of EXCFT values between ITU and ward admissions.

**Figure 5:**
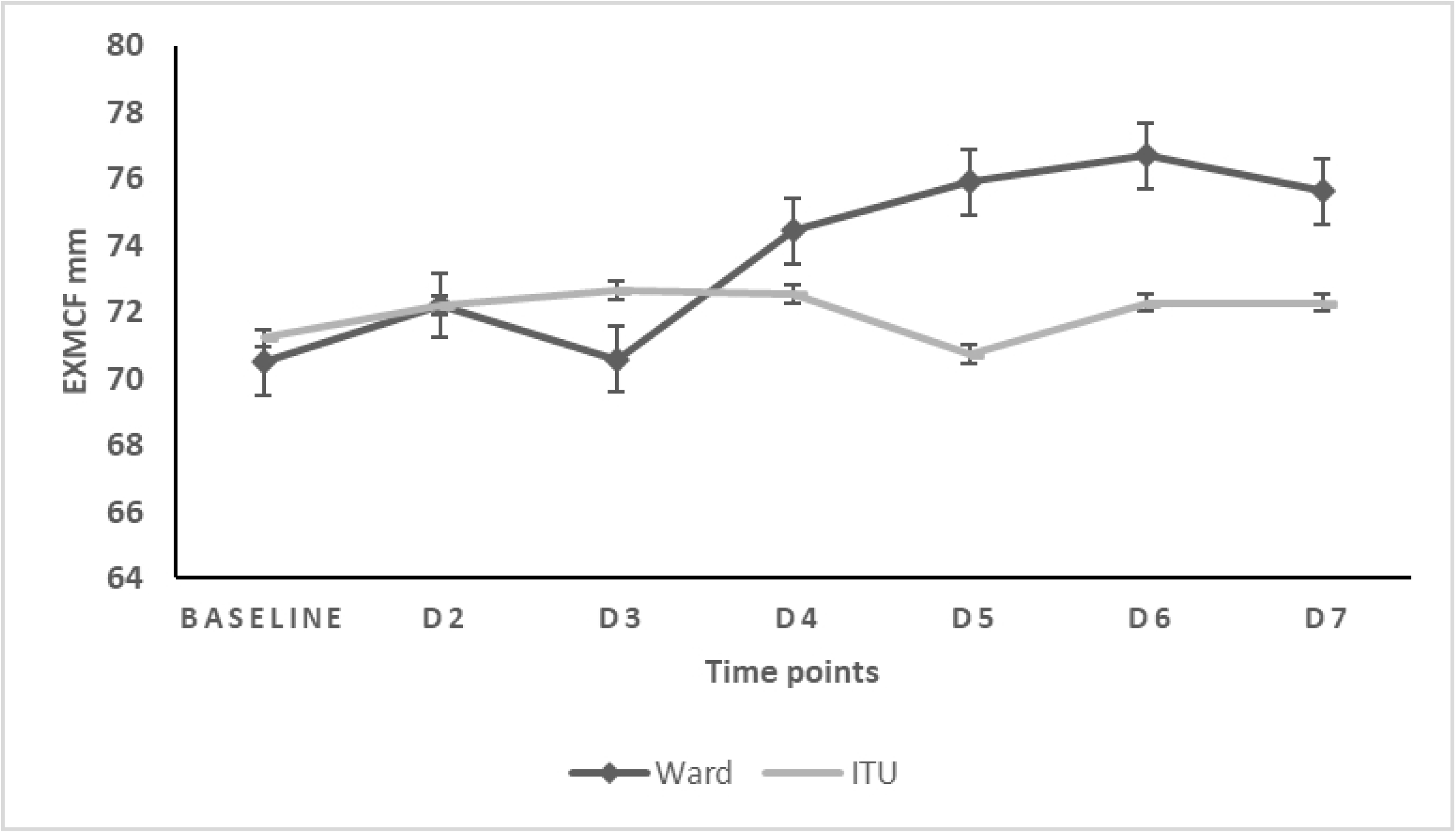
Values of EXMCF for COVID-19 patients over the first 7 days of hospital admission. Comparison of EXMCF values between ITU and ward patients.

**Figure 6:**
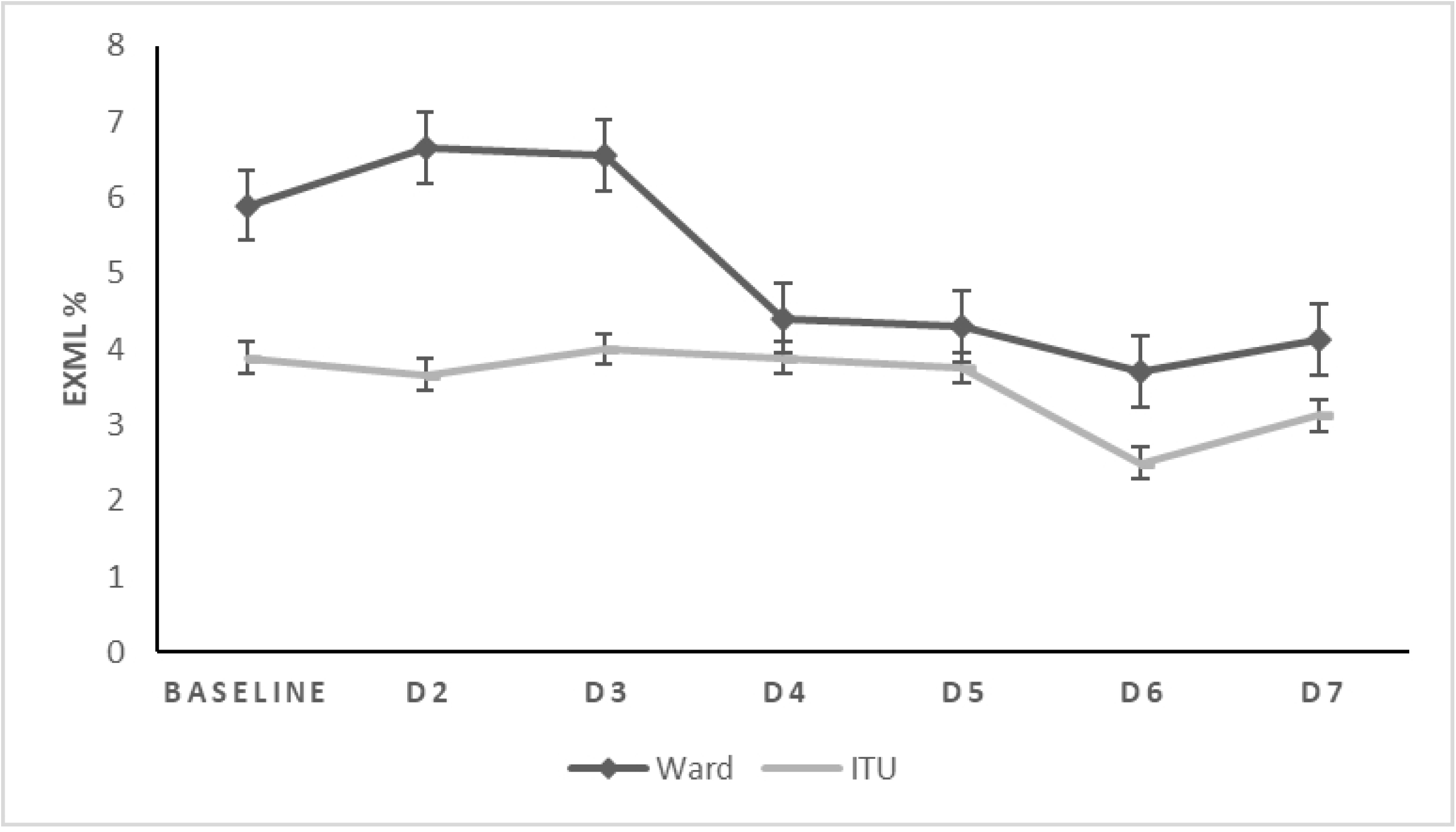
Values for EXML for COVID-19 patients over the first 7 days of hospital admission. Comparison of EXML values between ITU and ward patients.

**Figure 7:**
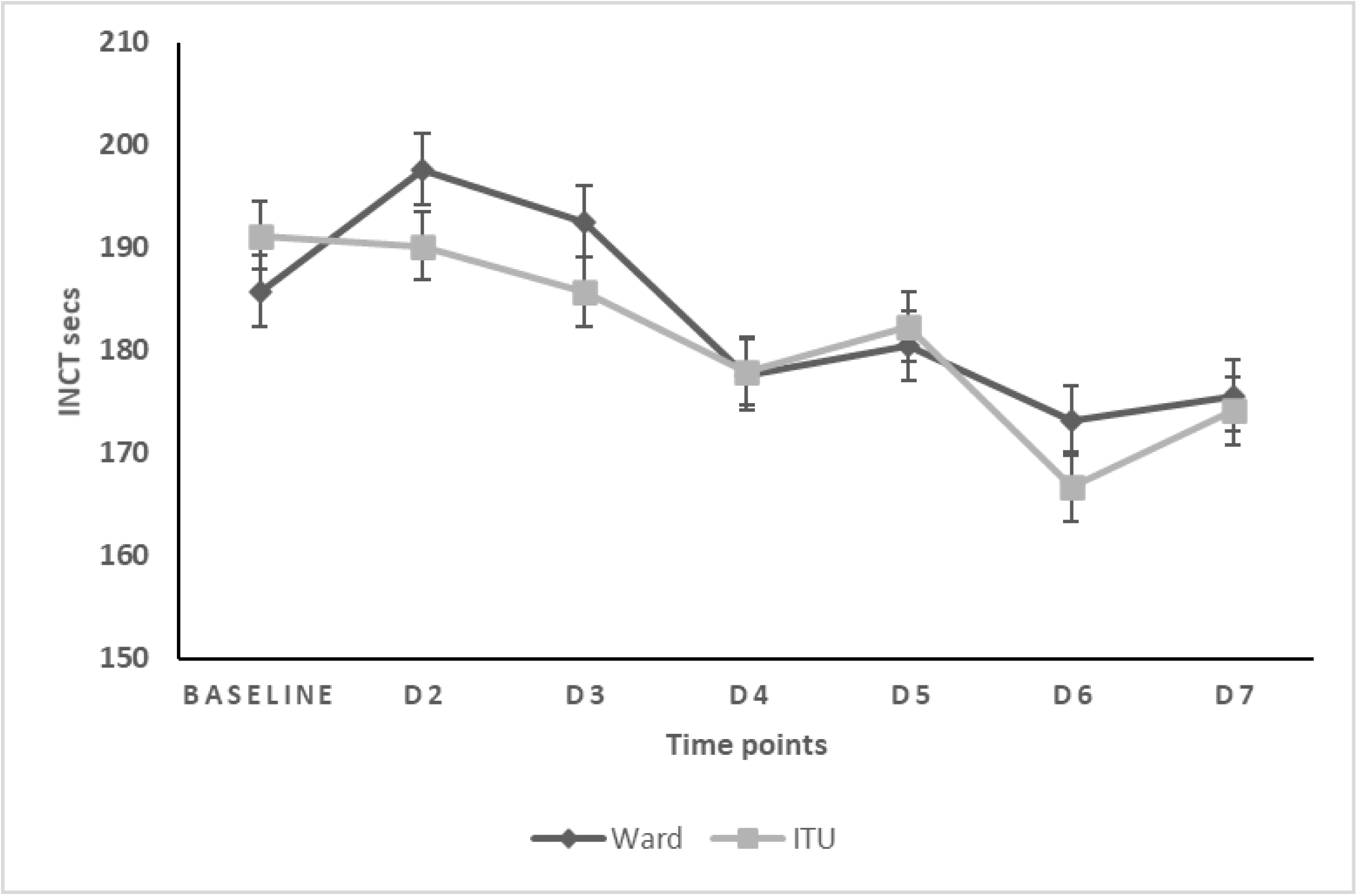
Values for INCT for COVID-19 patients over the first 7 days of hospital admission. Comparison of INCT values between ITU and ward patients.

**Figure 8:**
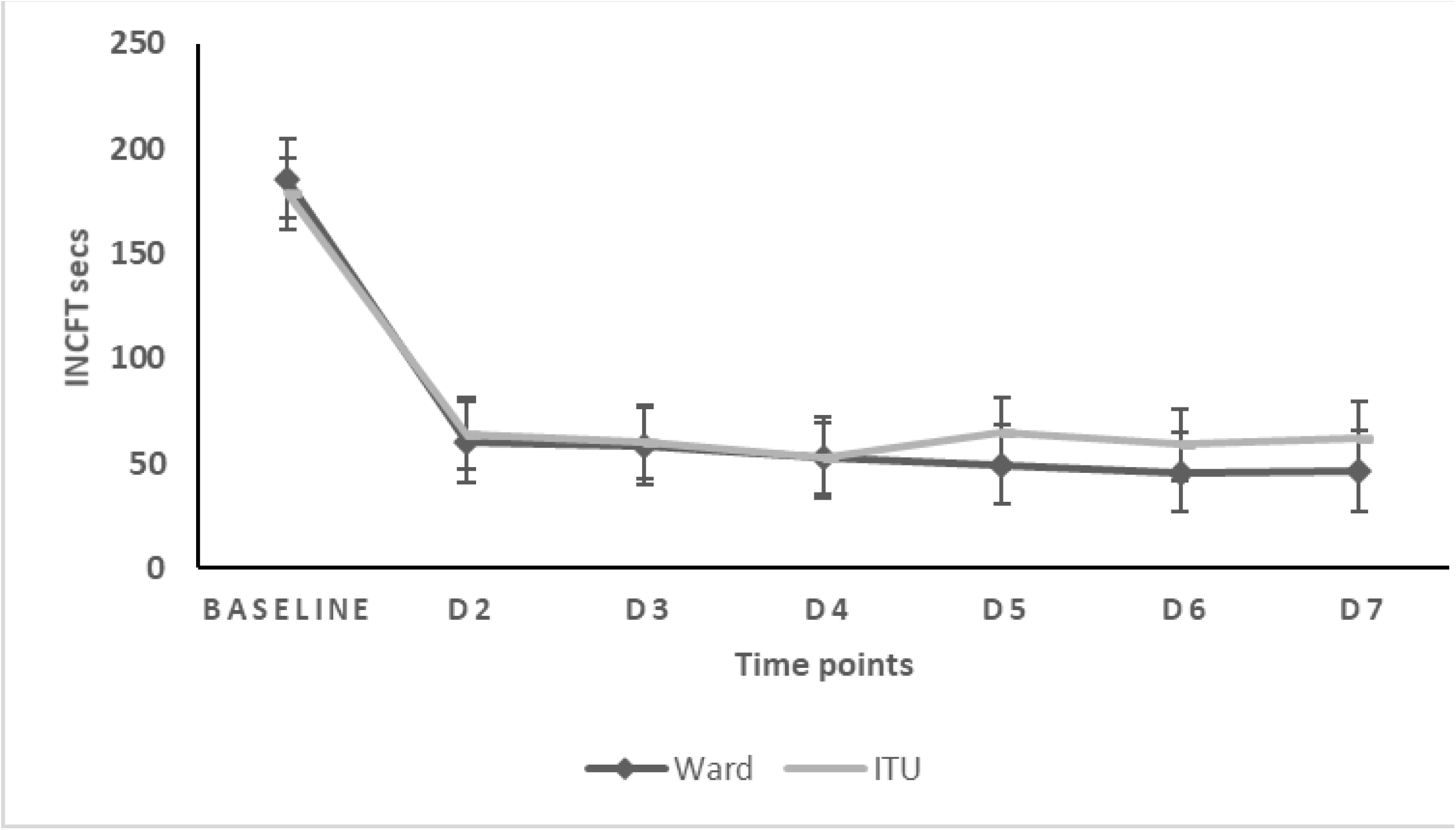
Values for INCFT for COVID-19 patients over the first 7 days of hospital admission. Comparison of INCFT values between ITU and ward patients.

**Figure 9:**
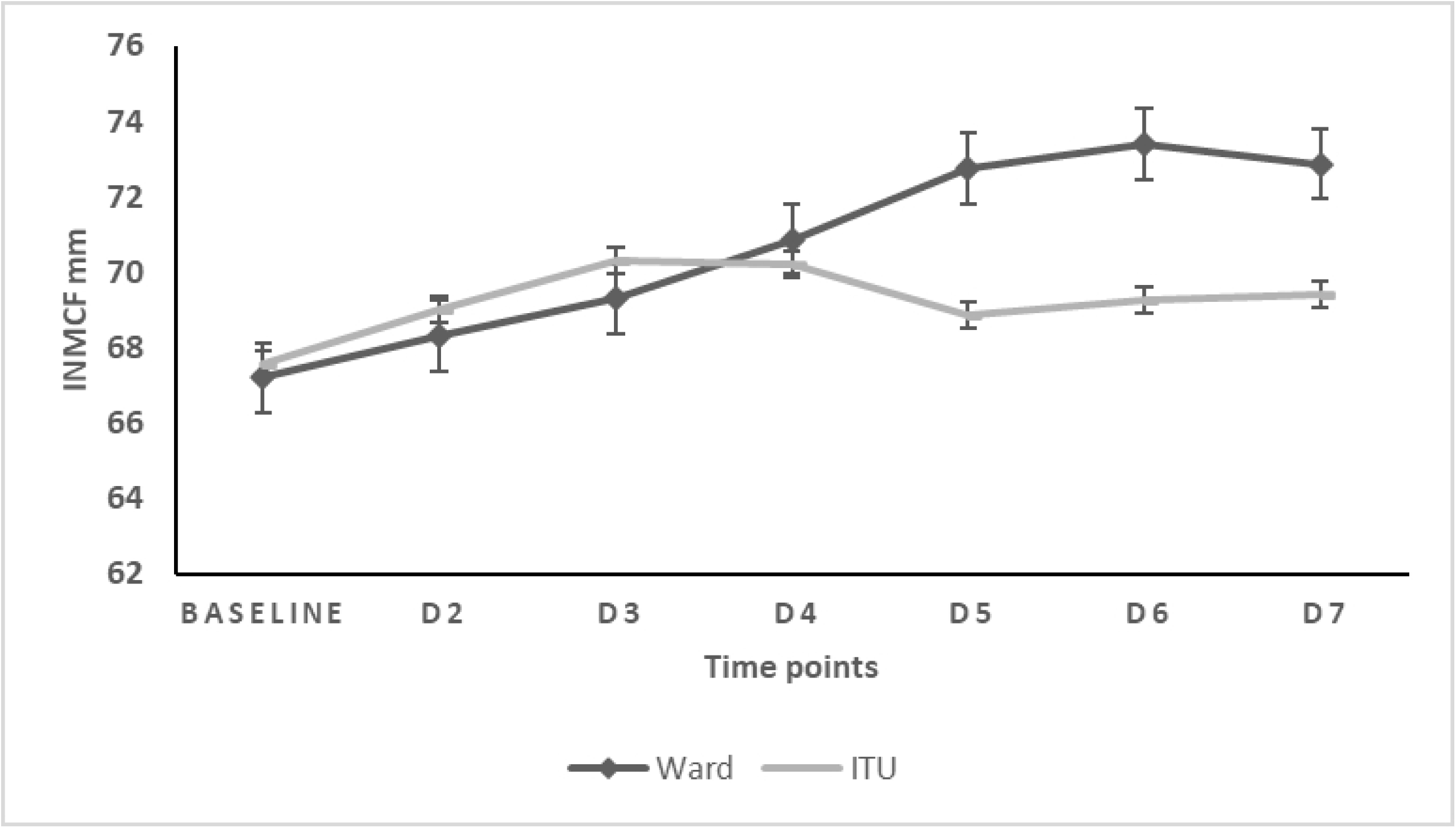
Values for INMCF for COVID-19 patients over the first 7 days of hospital admission. Comparison of INMCF values between the ITU and ward patients.

**Figure 10:**
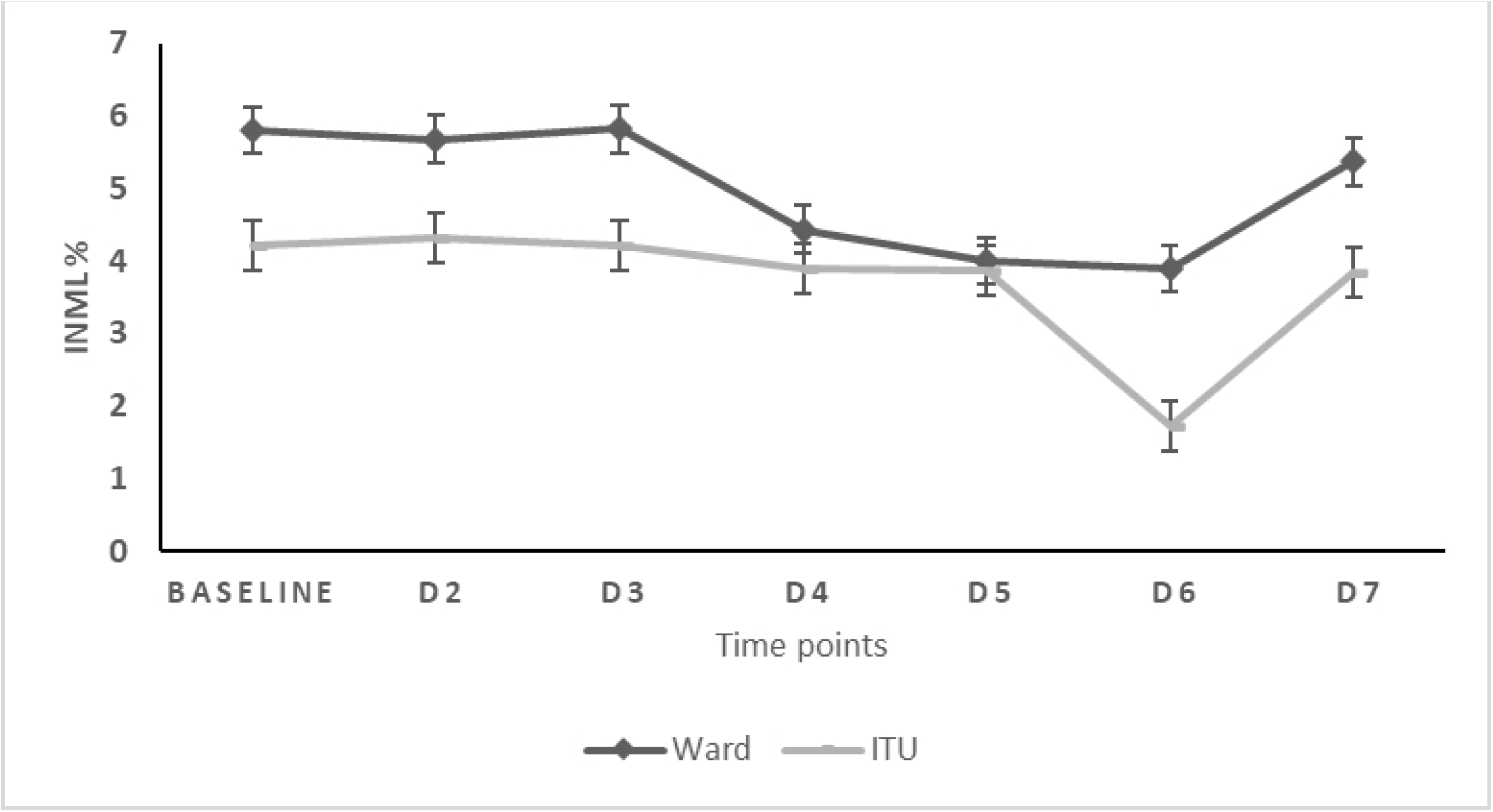
Values for INML for COVID-19 patients over the first 7 days of hospital admission. Comparison of INML values between ITU and ward patients.

## References

1. Li Q, Guan X, Wu P, Wang X, Zhou L, Tong Y, et al. Early Transmission Dynamics in Wuhan, China, of Novel Coronavirus–Infected Pneumonia. New England Journal of Medicine. 2020;382(13):1199–207.

2. Huang C, Wang Y, Li X, Ren L, Zhao J, Hu Y, et al. Clinical features of patients infected with 2019 novel coronavirus in Wuhan, China. The Lancet. 2020;395(10223):497–506.

3. Mehta P, McAuley DF, Brown M, Sanchez E, Tattersall RS, Manson JJ. COVID-19: consider cytokine storm syndromes and immunosuppression. Lancet. 2020;395(10229):1033–4.

4. Foley JH, Conway EM. Cross Talk Pathways Between Coagulation and Inflammation. Circ Res. 2016;118(9):1392–408.

5. O’Brien M. The reciprocal relationship between inflammation and coagulation. Topics in companion animal medicine. 2012;27(2):46–52.

6. D’Angelo G. Inflammation and coagulation: a “continuum” between coagulation activation and prothrombotic state. Journal of Blood Disorders. 2015;2.

7. Yin S, Huang M, Li D, Tang N. Difference of coagulation features between severe pneumonia induced by SARS-CoV2 and non-SARS-CoV2. J Thromb Thrombolysis. 2020:1–4.

8. Han H, Yang L, Liu R, Liu F, Wu KL, Li J, et al. Prominent changes in blood coagulation of patients with SARS-CoV-2 infection. Clinical chemistry and laboratory medicine. 2020.

9. Spiezia L, Boscolo A, Poletto F, Cerruti L, Tiberio I, Campello E, et al. COVID-19-Related Severe Hypercoagulability in Patients Admitted to Intensive Care Unit for Acute Respiratory Failure. Thrombosis and haemostasis. 2020.

10. Araya S, Mamo MA, Tsegay YG, Atlaw A, Aytenew A, Hordofa A, et al. Blood coagulation parameter abnormalities in hospitalized patients with confirmed COVID-19 in Ethiopia. PLoS One. 2021;16(6):e0252939.

11. Agbuduwe C, Basu S. Haematological manifestations of COVID-19: From cytopenia to coagulopathy. Eur J Haematol. 2020;105(5):540–6.

12. Tang N, Li D, Wang X, Sun Z. Abnormal coagulation parameters are associated with poor prognosis in patients with novel coronavirus pneumonia. Journal of thrombosis and haemostasis : JTH. 2020;18(4):844–7.

13. Naik BI, Pajewski TN, Bogdonoff DI, Zuo Z, Clark P, Terkawi AS, et al. Rotational thromboelastometry-guided blood product management in major spine surgery. J Neurosurg Spine. 2015;23(2):239–49.

14. Panigada M, Bottino N, Tagliabue P, Grasselli G, Novembrino C, Chantarangkul V, et al. Hypercoagulability of COVID-19 patients in Intensive Care Unit. A Report of Thromboelastography Findings and other Parameters of Hemostasis. Journal of thrombosis and haemostasis : JTH. 2020.

15. Ranucci M, Ballotta A, Di Dedda U, Bayshnikova E, Dei Poli M, Resta M, et al. The procoagulant pattern of patients with COVID-19 acute respiratory distress syndrome. Journal of Thrombosis and Haemostasis.n/a(n/a).

16. Hulshof A-M, Brüggemann RAG, Mulder MMG, van de Berg TW, Sels J-WEM, Olie RH, et al. Serial EXTEM, FIBTEM, and tPA Rotational Thromboelastometry Observations in the Maastricht Intensive Care COVID Cohort—Persistence of Hypercoagulability and Hypofibrinolysis Despite Anticoagulation. Frontiers in Cardiovascular Medicine. 2021;8(219).

17. Almskog LM, Wikman A, Svensson J, Wanecek M, Bottai M, van der Linden J, et al. Rotational thromboelastometry results are associated with care level in COVID-19. Journal of Thrombosis and Thrombolysis. 2021;51(2):437–45.

18. Raval JS, Burnett AE, Rollins-Raval MA, Griggs JR, Rosenbaum L, Nielsen ND, et al. Viscoelastic testing in COVID-19: a possible screening tool for severe disease? Transfusion. 2020;60(6):1131–2.

19. Boscolo A, Spiezia L, Correale C, Sella N, Pesenti E, Beghetto L, et al. Different Hypercoagulable Profiles in Patients with COVID-19 Admitted to the Internal Medicine Ward and the Intensive Care Unit. Thromb Haemost. 2020;120(10):1474–7.

20. Mitrovic M, Sabljic N, Cvetkovic Z, Pantic N, Zivkovic Dakic A, Bukumiric Z, et al. Rotational thromboelastometry (ROTEM) profiling of COVID–19 patients. Platelets. 2021;32(5):690–6.

21. Schenk B, Görlinger K, Treml B, Tauber H, Fries D, Niederwanger C, et al. A comparison of the new ROTEM®sigma with its predecessor, the ROTEMdelta. Anaesthesia. 2019;74(3):348–56.

22. Lier H, Vorweg M, Hanke A, Görlinger K. Thromboelastometry guided therapy of severe bleeding. Essener Runde algorithm. Hamostaseologie. 2013;33(1):51–61.

23. Crochemore T, Piza FMT, Rodrigues RDR, Guerra JCC, Ferraz LJR, Corrêa TD. A new era of thromboelastometry. Einstein (Sao Paulo). 2017;15(3):380–5.

24. Luis García de Guadiana R, Mulero MDR, Olivo MH, Rojas CR, Arenas VR, Morales MG, et al. Circulating levels of GDF-15 and calprotectin for prediction of in-hospital mortality in COVID-19 patients: A case series. J Infect. 2021;82(2):e40–e2.

25. Chen L, Long X, Xu Q, Tan J, Wang G, Cao Y, et al. Elevated serum levels of S100A8/A9 and HMGB1 at hospital admission are correlated with inferior clinical outcomes in COVID-19 patients. Cell Mol Immunol. 2020;17(9):992–4.

26. Shi H, Zuo Y, Yalavarthi S, Gockman K, Zuo M, Madison JA, et al. Neutrophil calprotectin identifies severe pulmonary disease in COVID-19. J Leukoc Biol. 2021;109(1):67–72.

27. Silvin A, Chapuis N, Dunsmore G, Goubet AG, Dubuisson A, Derosa L, et al. Elevated Calprotectin and Abnormal Myeloid Cell Subsets Discriminate Severe from Mild COVID-19. Cell. 2020;182(6):1401–18.e18.

28. Mahler M, Meroni P-L, Infantino M, Buhler KA, Fritzler MJ. Circulating Calprotectin as a Biomarker of COVID-19 Severity. Expert Rev Clin Immunol. 2021;17(5):431–43.

29. Engelmann B, Massberg S. Thrombosis as an intravascular effector of innate immunity. Nat Rev Immunol. 2013;13(1):34–45.

30. Sui J, Noubouossie DF, Gandotra S, Cao L. Elevated Plasma Fibrinogen Is Associated With Excessive Inflammation and Disease Severity in COVID-19 Patients. Front Cell Infect Microbiol. 2021;11:734005.

31. Gragnano F, Sperlongano S, Golia E, Natale F, Bianchi R, Crisci M, et al. The Role of von Willebrand Factor in Vascular Inflammation: From Pathogenesis to Targeted Therapy. Mediators of Inflammation. 2017;2017:5620314.

32. Chen J, Chung DW. Inflammation, von Willebrand factor, and ADAMTS13. Blood. 2018;132(2):141–7.

33. Tabatabai A, Rabin J, Menaker J, Madathil R, Galvagno S, Menne A, et al. Factor VIII and Functional Protein C Activity in Critically Ill Patients With Coronavirus Disease 2019: A Case Series. A A Pract. 2020;14(7):e01236.

34. Rauch A, Labreuche J, Lassalle F, Goutay J, Caplan M, Charbonnier L, et al. Coagulation biomarkers are independent predictors of increased oxygen requirements in COVID-19. Journal of Thrombosis and Haemostasis. 2020;18(11):2942–53.

35. Zhang Y, Cao W, Jiang W, Xiao M, Li Y, Tang N, et al. Profile of natural anticoagulant, coagulant factor and anti-phospholipid antibody in critically ill COVID-19 patients. Journal of thrombosis and thrombolysis. 2020;50(3):580–6.

36. Goshua G, Pine AB, Meizlish ML, Chang C-H, Zhang H, Bahel P, et al. Endotheliopathy in COVID-19-associated coagulopathy: evidence from a single-centre, cross-sectional study. The Lancet Haematology. 2020;7(8):e575–e82.

37. Ward SE, Curley GF, Lavin M, Fogarty H, Karampini E, McEvoy NL, et al. Von Willebrand factor propeptide in severe coronavirus disease 2019 (COVID-19): evidence of acute and sustained endothelial cell activation. British journal of haematology. 2021;192(4):714–9.

38. Zakai NA, McClure LA. Racial differences in venous thromboembolism. J Thromb Haemost. 2011;9(10):1877–82.

